# A new combination testing methodology to identify accurate and economical point-of-care testing strategies

**DOI:** 10.1101/2021.06.15.21257351

**Authors:** Sanjay Jain, Jónas Oddur Jónasson, Jean Pauphilet, Barnaby Flower, Maya Moshe, Gianluca Fontana, Sutharsan Satkunarajah, Richard Tedder, Myra McClure, Hutan Ashrafian, Paul Elliott, Wendy S Barclay, Christina Atchison, Helen Ward, Graham Cooke, Ara Darzi, Kamalini Ramdas

## Abstract

**Background:** Quick, cheap and accurate point-of-care testing is urgently needed to enable frequent, large-scale testing to contain COVID-19. Lateral flow tests for antigen and antibody detection are an obvious candidate for use in community-wide testing, because they are quick and cheap relative to lab-processed tests. However, their low accuracy has limited their adoption. We develop a new methodology to increase the diagnostic accuracy of a combination of cheap, quick and inaccurate index tests with correlated or discordant outcomes, and illustrate its performance on commercially available lateral flow immunoassays (LFIAs) for Sars-CoV-2 antibody detection.

**Methods and Findings:** We analyze laboratory test outcomes of 300 serum samples from health care workers detected with PCR-confirmed SARS-Cov-2 infection at least 21 days prior to sample collection, and 500 pre-pandemic serum samples, from a national seroprevalence survey, tested using eight LFIAs (Abbott, Biosure/Mologic, Orientgene-Menarini, Fortress, Biopanda I, Biopanda II, SureScreen and Wondfo) and Hybrid DABA as reference test. For each of 14 two-test combinations (e.g., Abbott, Fortress) and 16 three-test combinations (e.g., Abbott, Fortress, Biosure/Mologic) used on at least 100 positive and 100 negative samples, we classify an outcome sequence – e.g., (+,–) for (Abbott, Fortress) – as positive if its combination positive predictive value (CPPV) exceeds a given threshold, set between 0 and 1. Our main outcome measures are the sensitivity and specificity of different classification rules for classifying the outcomes of a combination test. We define testing possibility frontiers which represent sensitivity and false positive rates for different thresholds. The envelope of frontiers further enables test selection.

The eight index tests individually meet neither the UK Medicines and Healthcare Products Regulatory Agency’s 98% sensitivity and 98% specificity criterion, nor the US Center for Disease Control’s 99.5% specificity criterion. Among these eight tests, the highest single-test LFIA specificity is 99.4% (with a sensitivity of 65.2%) and the highest single-test LFIA sensitivity is 93.4% (with a specificity of 97.4%). Using our methodology, a two-test combination meets the UK Medicines and Healthcare Products Regulatory Agency’s criterion, achieving sensitivity of 98.4% and specificity of 98.0%. While two-test combinations meeting the US Center for Disease Control’s 99.5% specificity criterion have sensitivity below 83.6%, a three-test combination delivers a specificity of 99.6% and a sensitivity of 95.8%.

**Conclusions:** Current CDC guidelines suggest combining tests, noting that “performance of orthogonal testing algorithms has not been systematically evaluated” and highlighting discordant outcomes. Our methodology combines available LFIAs to meet desired accuracy criteria, by identifying testing possibility frontiers which encompass benchmarks, enabling cost savings. Our methodology applies equally to antigen testing and can greatly expand testing capacity through combining less accurate tests, especially for use cases needing quick, accurate tests, e.g., entry to public spaces such as airports, nursing homes or hospitals.

## INTRODUCTION

Quick, cheap and accurate point-of-care (POC) antigen and antibody tests are urgently needed to enable frequent, large-scale testing to contain COVID-19.[1–3] Accurate Rapid Antigen Tests could allow for significant scale-up of the frequency and scope of diagnostic testing [4–6] and enable quick and accurate testing in use cases such as entry to public spaces including airports, nursing homes and hospitals. Accurate POC antibody tests will allow for large-scale serological surveys to estimate prevalence,[7] identify individuals who receive false negative RT-PCR results [8] or have high quality convalescent plasma, and enable redeployment of recovered individuals into the community.[9] Yet, the accuracy of existing lateral flow antigen and antibody tests is low.

We contribute a new and universal methodology to improve diagnostic accuracy, which relies on using multiple index tests, and we illustrate this methodology in the context of Covid-19 POC antibody tests.

For POC antibody detection, lateral flow immunoassays (LFIAs) are an obvious candidate for use in community-wide testing, because they are quick and cheap relative to lab-processed antibody tests (e.g., ELISAs). However, their low accuracy has limited their adoption.[1,10–12] Many LFIAs meeting the US Center for Disease Control’s (CDC) 99.5% specificity criterion have low sensitivity. Similarly, few LFIAs have won approval by meeting the UK Medicines and Healthcare Products Regulatory Agency’s (MHRA) 98% sensitivity and 98% specificity criteria.[13]

Combination testing – i.e., assessing each sample using more than one test – can in theory increase the accuracy of tests whose outcomes are independent. [14] The CDC’s current guidelines for COVID-19 antibody testing suggests combining tests, stating that *“the performance of orthogonal testing algorithms has not been systematically evaluated but can be estimated using an online calculator from the FDA”*,^1^ which assumes independent test outcomes. The FDA guidelines highlight that results can be “discordant” biologically– for instance when the antigens (e.g., spike protein or nucleocapsid protein) or the Ig classes (total Ig, IgG, or IgM) that are detected in two separate tests are different. There are no guidelines on how to handle correlated or discordant results. However, in practice, outcomes of tests of different formats which detect the same antibodies or use the same substrate are likely to be correlated but not identical, generating discordant results.

We develop a new methodology to classify the outcomes of combination tests with correlated or discordant outcomes. We illustrate, using commercially available LFIAs, that our methodology increases diagnostic accuracy and the policy maker’s choices, relative to existing heuristics.

Our methodology enables identification of LFIA combination tests – and appropriate classification rules – that meet policy makers’ accuracy criteria where individual tests might not, at a cost that opens up a much-needed pathway to large-scale and frequent antibody testing.

Any combination of tests requires a method to classify each *outcome sequence*—e.g., (+, +) or (+, -) for a particular two-test combination—as positive or negative. Existing methods include the ‘*majority*’, ‘believe the positive’ (i.e., ‘*any’*), and ‘believe the negative’ (i.e., ‘*all’*) heuristic rules, or in some cases additional arbitrator tests.[15–21] Instead, we classify each outcome sequence based on its *combination positive predictive value*, and identify the *testing possibility frontier*, i.e., the set of achievable sensitivity and specificity parameters for a combination test.

We compare this method to existing benchmarks using data from a national seroprevalence study.[13] Our methodology complements prior literature on the utility of cheap and less accurate tests [22,23]and simple combination testing, which assumes classification heuristics such as *any, all* or *majority*. [19–21]

## METHODS

### Study Sample

Our study sample is 300 antibody positive and 500 pre-pandemic antibody negative serum samples from a REACT-2 study, which evaluated the accuracy and suitability of commercially available SARS-CoV-2 antibody LFIAs for a national random population sampled seroprevalence survey.[13] We consider eight index tests (see Table 1, Column 2), each performed on a subset of our study sample.

**Table 1:**
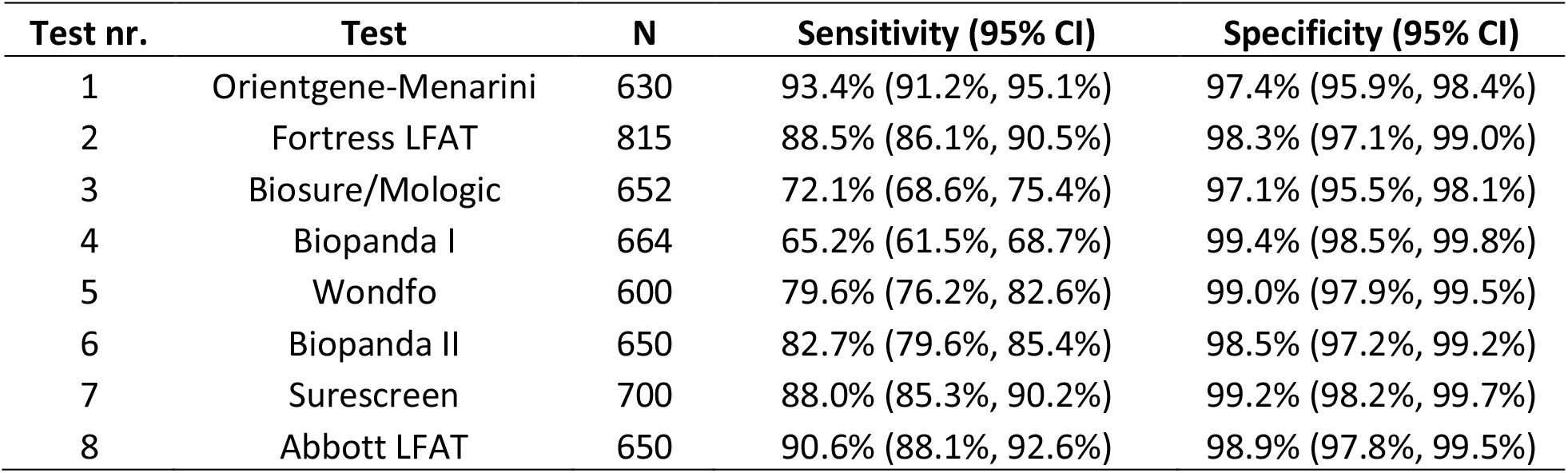
A list of our eight index tests and their testing accuracy.

| Test nr. | Test | N | Sensitivity (95% CI) | Specificity (95% CI) |
| --- | --- | --- | --- | --- |
| 1 | Orientgene-Menarini | 630 | 93.4% (91.2%, 95.1%) | 97.4% (95.9%, 98.4%) |
| 2 | Fortress LFAT | 815 | 88.5% (86.1%, 90.5%) | 98.3% (97.1%, 99.0%) |
| 3 | Biosure/Mologic | 652 | 72.1% (68.6%, 75.4%) | 97.1% (95.5%, 98.1%) |
| 4 | Biopanda I | 664 | 65.2% (61.5%, 68.7%) | 99.4% (98.5%, 99.8%) |
| 5 | Wondfo | 600 | 79.6% (76.2%, 82.6%) | 99.0% (97.9%, 99.5%) |
| 6 | Biopanda II | 650 | 82.7% (79.6%, 85.4%) | 98.5% (97.2%, 99.2%) |
| 7 | Surescreen | 700 | 88.0% (85.3%, 90.2%) | 99.2% (98.2%, 99.7%) |
| 8 | Abbott LFAT | 650 | 90.6% (88.1%, 92.6%) | 98.9% (97.8%, 99.5%) |

The antibody positive samples were collected in May 2020, from adult National Health Service (NHS) workers who tested positive for SARS-CoV-2 through a nasopharyngeal swab PCR test, or showed symptoms, at least 21 days prior and had not been hospitalized, using hybrid DABA (hybrid spike protein receptor binding domain double antigen binding assay detecting antibody to RBD) as reference test. The antibody negative samples are from 2018, collected during the Airwaves study.[24] Sample sizes were determined in the prior study.[13] The REACT-2 study had significant patient and public input. We will disseminate results of our study to participants, once published.

Index tests were performed by laboratory technicians blind to the participants’ underlying condition, according to manufacturers’ instructions, and the result scored as either positive or negative for IgG.

Since the data was originally collected to evaluate the diagnostic performance of each index test, the data collection was not designed to prioritize applying each test to each sample. As a result, our analysis relies on fewer observations for each combination of tests. Specifically, we consider two-test and three-test combinations whose index tests were each evaluated on at least 100 antibody positive and at least 100 antibody negative samples. More observations for each combination of tests (e.g., through prospective data collection) would result in smaller confidence intervals for CPPVs and combination testing sensitivities and specificities (reported in Supplementary information sections 2 and 3).

### Individual Test Performance

We calculate the sensitivity and specificity of each index test against the DABA reference test and 95% confidence intervals using the Wilson method.[25] Supplementary information section 1 includes the pairwise correlation of outcomes (as well as true positives, false positives, true negatives, and false negatives).

### Combination Test Performance

Applying two (three) index tests to the same subject can result in four (eight) outcome sequences, e.g., (+,+), (+, –), (–, +), or (–, –) for a two-test combination. A useful combination test methodology must classify each outcome sequence as either a positive or a negative outcome, in a data-driven way that leverages the fact that some of the index tests are more accurate than others and also accounting for correlations amongst the test outcomes.

Given any combination test, our methodology classifies a sample as either positive or negative, depending on the outcome sequence obtained (see Figure 1). A combination of two (three) tests has four (eight) possible outcome sequences. This simple, data-driven 4-step classification methodology leverages the inherent relative accuracies of the index tests and accounts for correlations amongst the test outcomes.

**Figure 1:**
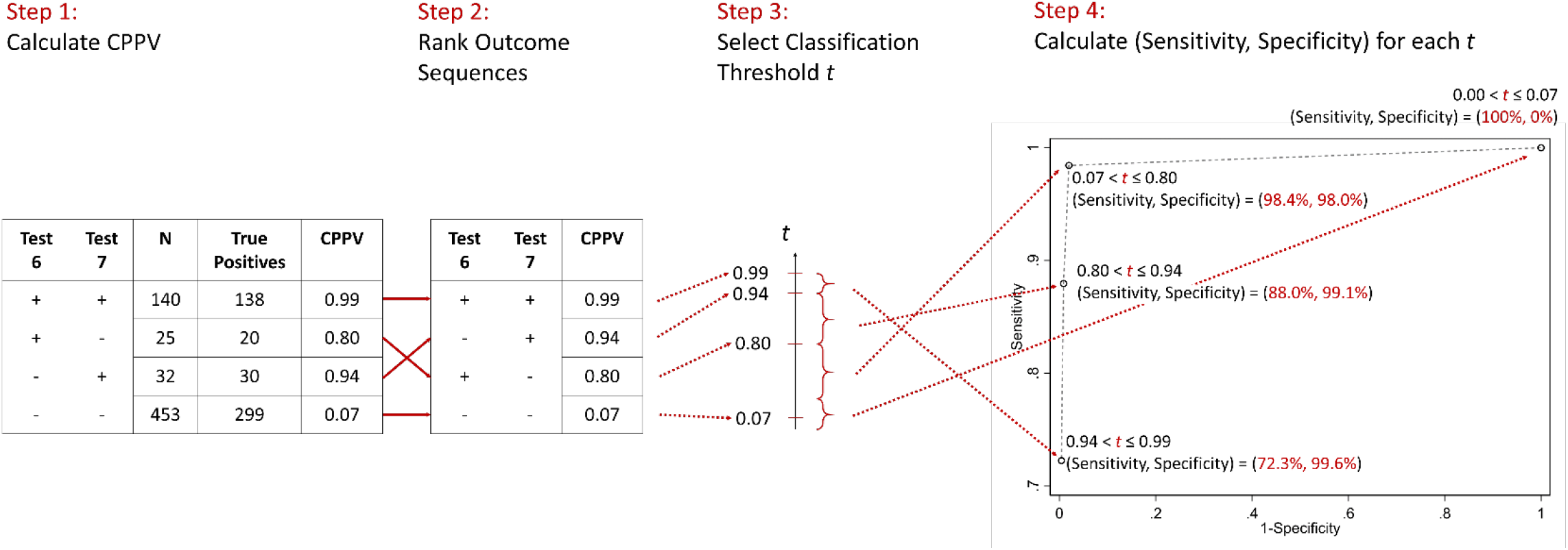
Illustrative example of the four-step classification method for combining tests 6 and 7.

**Step 1**. Extending the concept of PPV of a single test, we calculate the *combination positive predictive value* (CPPV) for each outcome sequence as the number of cases with that outcome sequence that are condition-positive, divided by the number that test positive. Supplementary information sections 2 and 3 include the CPPVs of all outcome sequences for all combinations of two or three tests along with confidence intervals.

**Step 2**. We rank outcome sequences by their CPPV. The highest-CPPV outcome sequence has the highest proportion of true positives.

**Step 3**. We use a CPPV threshold between 0 and 1 to classify each outcome sequence, with outcome sequences with CPPVs above or equal to the threshold classified as positive and the rest as negative.

**Step 4**. For each threshold value, we calculate the ‘implied’ sensitivity and specificity of the combination test in the traditional manner. A higher (lower number?) threshold will emphasize specificity (sensitivity). We represent the distinct (sensitivity, specificity) possibilities for each combination test through a *testing possibility frontier*, which represents the maximum achievable sensitivity (specificity) for a given level of specificity (sensitivity). Points on the frontier *dominate* those within it on at least one of these two dimensions.

This methodology has three main advantages. First, it enables flexibly choosing a threshold for a given combination of tests, to emphasize high sensitivity, high specificity, or both. It also generates an *envelope* of frontiers of multiple combination tests, enabling even greater flexibility in test selection. Second, it combines tests with different predictive accuracy in a data-driven way. By ranking outcome sequences by CPPV, it essentially places more weight on more accurate index tests, when classifying outcome sequences as positive or negative. Third, unlike the formula provided on the CDC website as of September 25 2020 which gives the maximum possible sensitivity advantage for a combination test^2^ assuming independent outcomes, we develop data-driven implied sensitivity and specificity estimates that reflect the inherent correlations. High correlation results in minimal improvement in sensitivity/specificity over the index tests.

### Benchmark Heuristics for Combination Testing

We compared the accuracy obtained using our threshold rule with that of three common benchmark heuristics.[18]

**The ANY heuristic** classifies an outcome sequence as positive if any of its index tests returns a positive outcome.

**The ALL heuristic** classifies an outcome sequence as positive only if all of its index tests return a positive outcome.

**The MAJORITY heuristic** classifies an outcome sequence as positive only if the majority of its index tests return a positive outcome.

These benchmark heuristics have two significant drawbacks relative to the threshold rule. First, they treat all tests in a combination as equal, regardless of their predictive accuracy. Thus applying the MAJORITY heuristic to a combination test with one accurate and two inaccurate index tests will worsen accuracy. In contrast, our methodology assigns greater weight to tests with higher predictive accuracy. Second, each benchmark heuristic results in a single set of sensitivity and specificity values. By design, the ANY heuristic often results in high sensitivity (and low specificity) and the ALL heuristic in high specificity (and low sensitivity). In contrast, our methodology offers the policy maker a menu of attainable sensitivity-specificity values given any combination of index tests, so that the same tests can be used for populations with different accuracy needs.

## RESULTS

### Individual Test Performance

The sensitivity and specificity of the eight index tests using DABA as reference test are reported in Table 1. Specificity is generally high (97.1% to 99.4%) and sensitivity low (65.2% to 93.4%). Moreover, the highest specificity (sensitivity) index test had the lowest sensitivity (specificity). No single test meets the CDC or MHRA criteria. Test outcomes are highly, but not perfectly, correlated (average pairwise correlation: 0.75, range: 0.57 to 0.89).

### Two-Test Combination Performance

Figure 1 illustrates our four step methodology, using a combination of two index tests – test 6 (Biopanda II) and test 7 (Surescreen). Each of the four possible outcome sequences (+, +), (+, –), (–, +), and (–, –) provides a threshold value that generates a combination test strategy with distinct sensitivity and specificity. Ignoring the lowest threshold value (which trivially classifies all samples as positive) we observe that combining these two tests can achieve a testing strategy with (sensitivity, specificity) values ranging from (98.4% [CI: 97.2%-99.1%], 98.0% [CI: 96.7%-98.9%]) to (72.3% [CI: 68.7%-75.6%], 99.6% [CI: 98.7%-99.9%]) in our data. The former meets MHRA’s but not the CDC’s criteria, while the latter meets the CDC’s criteria albeit with very low sensitivity, but not the MHRA’s. In particular, a threshold between 0.01 and 0.80 classifies outcome sequences in which either test returns a positive outcome (corresponding to the ANY heuristic) as positive, for a sensitivity of 98.4% [CI: 97.2%-99.1%] (given index test sensitivities of 82.7% [CI: 79.6%-85.4%] and 88.0% [CI: 85.3%-90.2%]), with minimal reduction in specificity.

Figure 2 (top) plots the testing possibility frontiers for the 16 two-test combinations in which each index test was used on at least 100 antibody negative and at least 100 antibody positive samples (all estimates and their confidence intervals are included in Supplementary information section 2). Each marked point corresponds to a threshold value for a particular combination test. For ease of viewing, we ignore thresholds which result in sensitivity or specificity below 50%. To gain intuition, in Figure 2 (bottom) we include the results of the ANY and ALL heuristics (the MAJORITY heuristic requires three or more tests) for the same 16 two-test combinations. The marker points with highest sensitivity generally correspond to the ANY heuristic and those with highest specificity to the ALL heuristic. No benchmark improves on a strategy obtained through our methodology.

**Figure 2:**
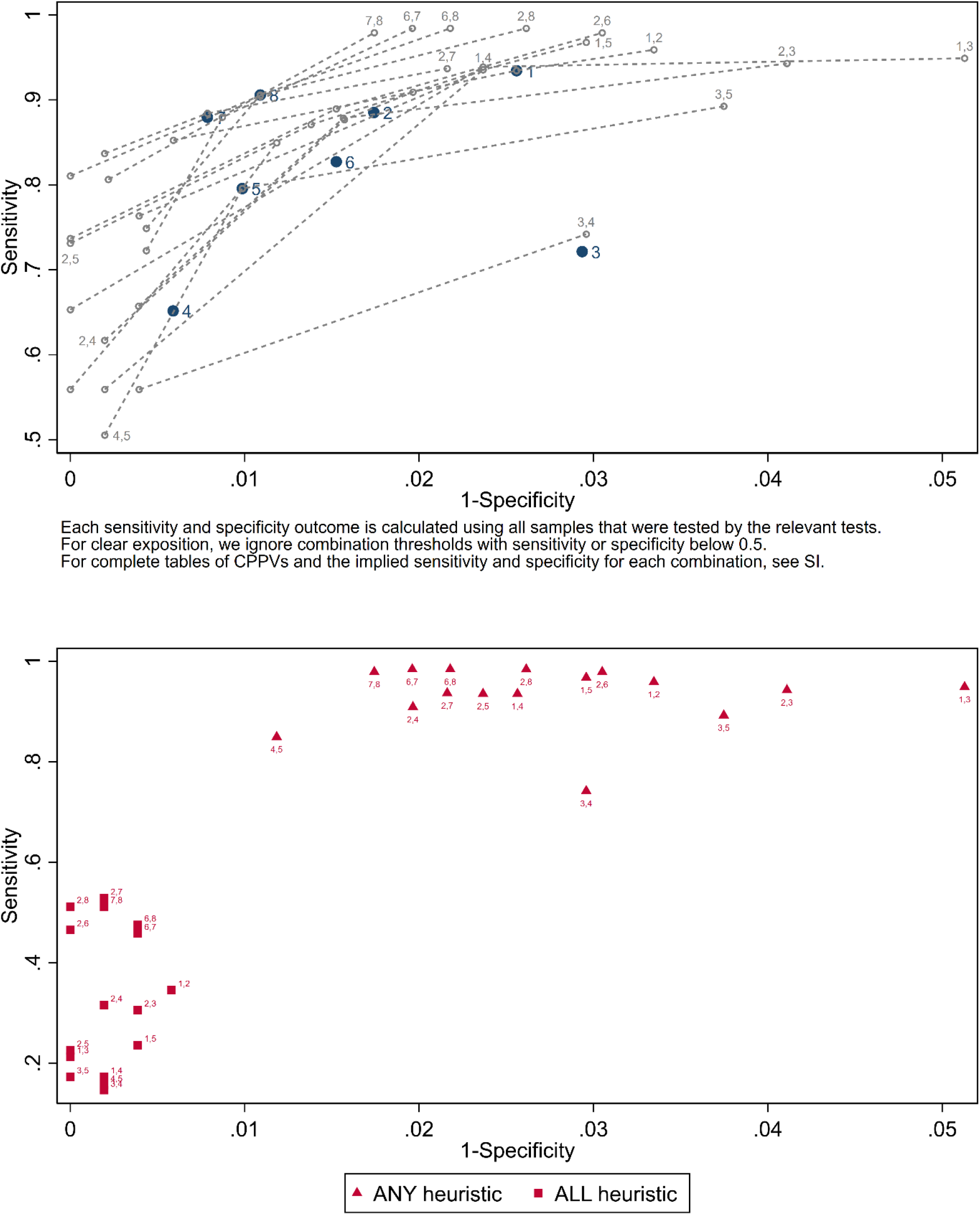
Two test combination performance.

Figure 2 also shows that (6, 7) – i.e., (Biopanda II, Surescreen) – is the only 2-test combination that meets the MHRA criteria, while (2, 8) and (6, 8) – i.e., (Fortress, Abbott) and (Biopanda II, Abbott) – achieve an estimated sensitivity above 98% and specificity above only 96%. Our point estimates indicate that the CDC’s 99.5% specificity criterion is met by 15 out of the 16 two-test combinations, for high-enough thresholds (see full details in Supplementary information section 2). Only outcome sequences with suitably high CPPVs are classified as positive in this case. However, the resulting sensitivities are very low: the highest sensitivity achievable while meeting the CDC criteria is 83.7% [CI: 80.8%-86.2%], for the combination test (2, 7), i.e., (Fortress, Surescreen).

### Three-Test Combination Performance

For illustration, we consider test 6 (Biopanda II) and test 7 (Surescreen) as before, adding test 8 (Abbott) to the combination. Three tests result in eight possible outcomes sequences: (+, +, +), (+, +, –), (+, –, +), (+, –, –), (–, +, +), (–, +, –), (–, –, +), or (–, –, –), and therefore eight threshold values. Note that in our approach the ranking of CPPVs is subject to sampling error. Larger sample size, as well as robust optimization techniques[26] can be used to account for this issue.

Figure 3 provides an overview of the four steps and Figure 4 displays the testing possibility frontier for these three tests. Sensitivity/specificity values ranging from (100.0% [CI: 99.4%-100%], 97.4% [CI: 95.9%-98.4%]) to (80.6% [CI: 77.4%-83.5%], 99.8% [CI: 99.0%-100%]) can be obtained, depending on the threshold. A threshold value of 0.6 maximizes sensitivity while maintaining a specificity estimate above 98.0%, resulting in a sensitivity/specificity of (96.3% [CI: 94.6%-97.5%], 98.9% [CI: 97.8%-99.5%]), classifying 5 outcome sequences, (+, +, +), (+, +, –), (+, –, +), (–, +, +) and (–, +, –), as positive and the remaining 3 as negative. Figure 4 shows that for this three-test combination, no benchmark heuristic (ANY/ALL/MAJORITY) achieves higher sensitivity than 94.8% and specificity above 98%. The threshold rule achieves higher sensitivity because at a threshold of 0.6, it classifies the outcome sequence (–, +, –) as positive, despite two negative index tests.

**Figure 3:**
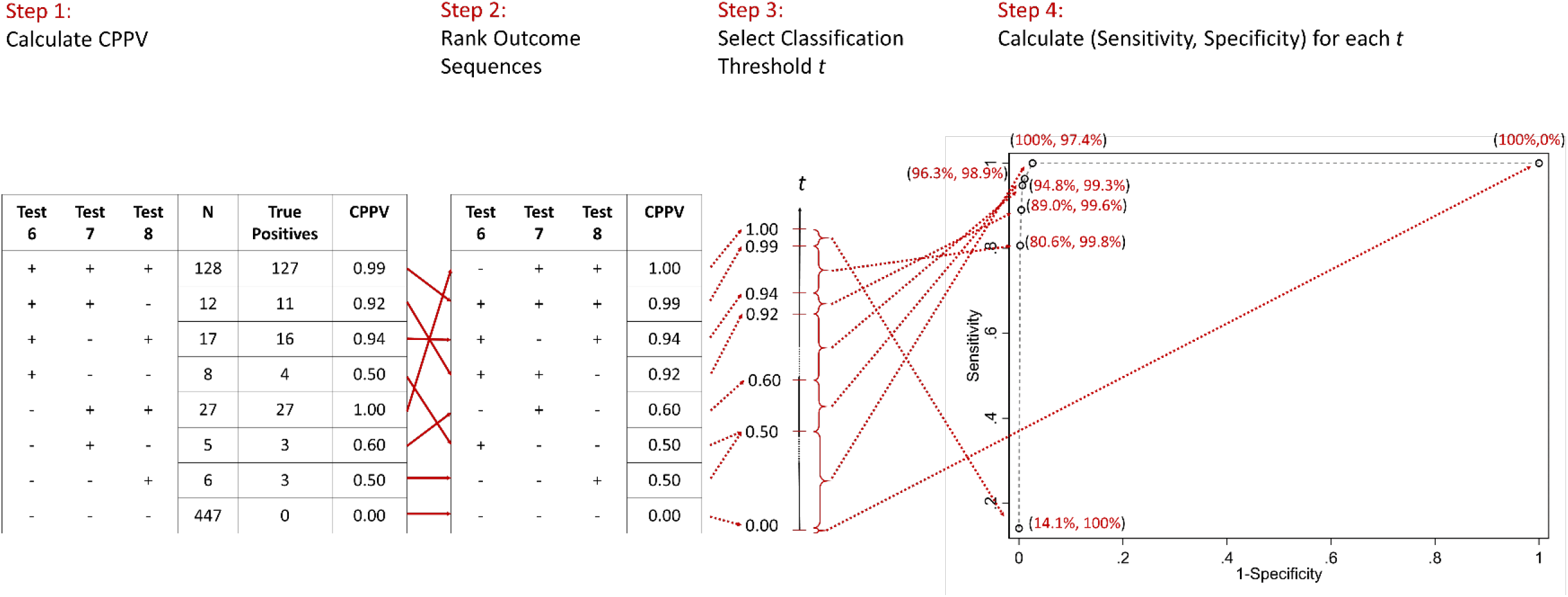
Illustrative example of the four-step classification method for combining tests 6, 7, and 8.

**Figure 4:**
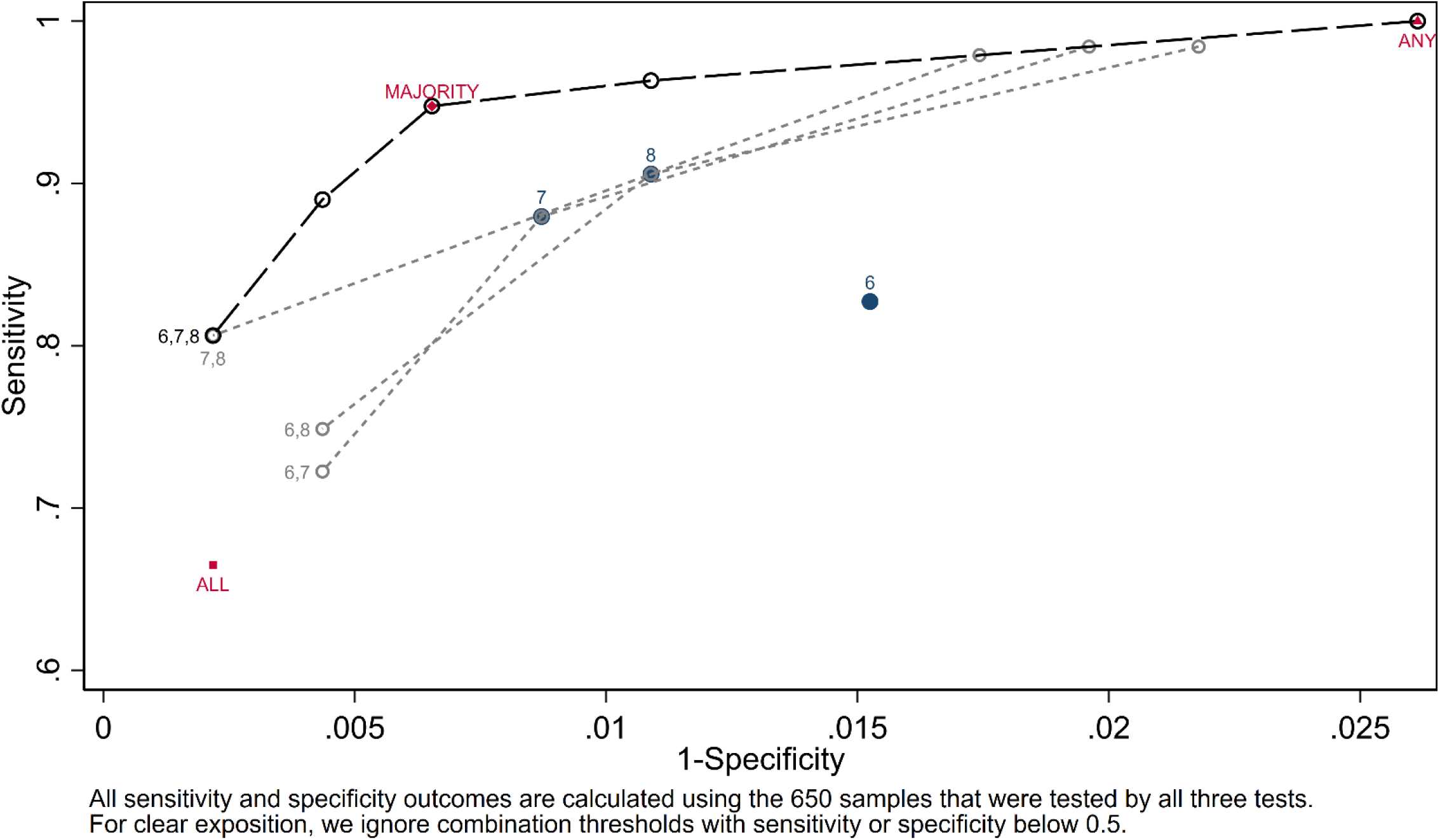
Performance comparison for combining tests 6, 7, and 8.

Figure 5 presents the testing possibility frontiers for the 14 three-test combinations that satisfy our inclusion criteria on sample size for the index tests (all estimates and their confidence intervals are included in Supplementary information section 2). Again, the marked points correspond to distinct threshold values on each frontier and we ignore threshold values which result in sensitivity or specificity below 50%.

**Figure 5:**
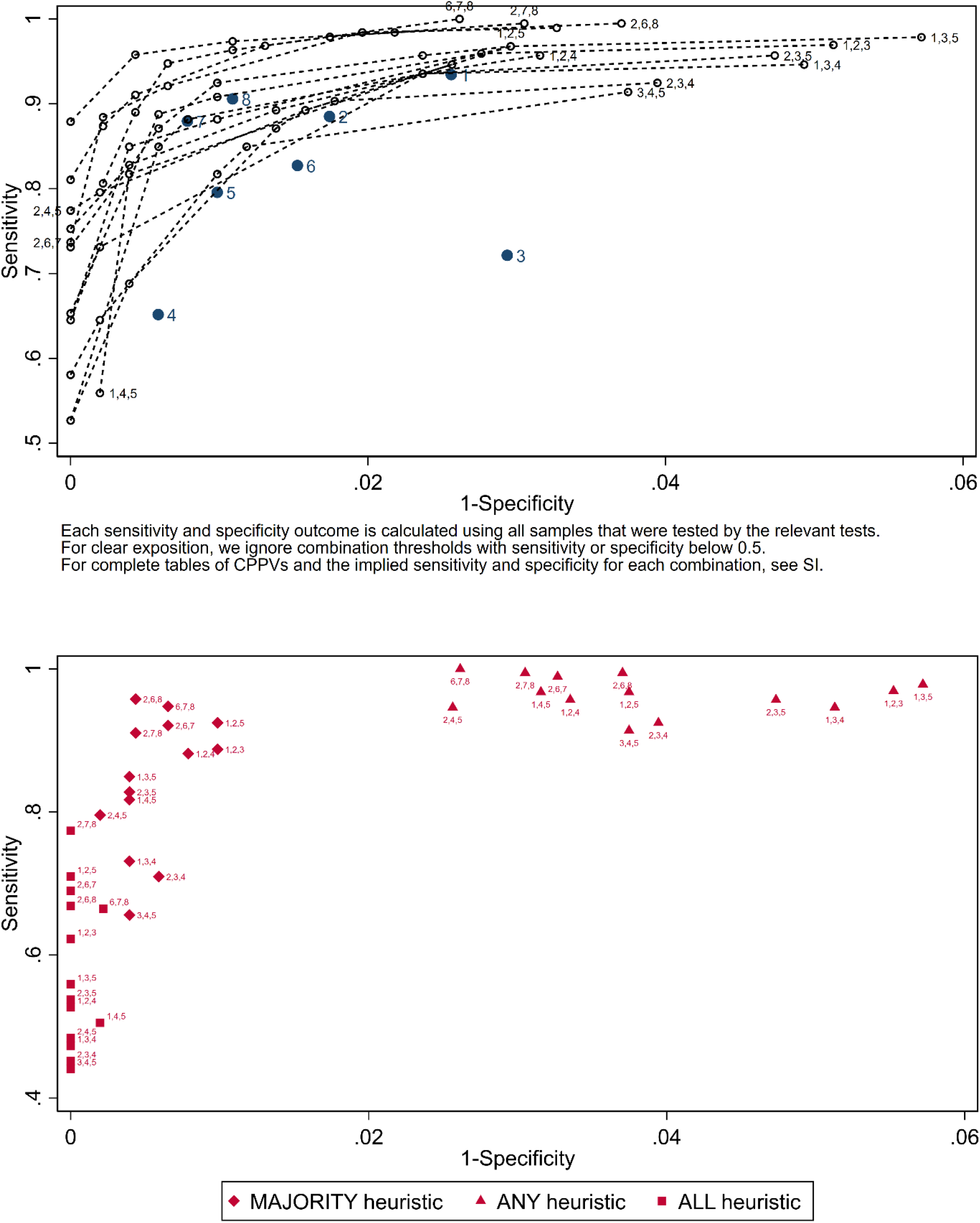
Three test combination performance.

Only one three-test combination – of index tests 2 (Fortress), 6 (Biopanda II), and 7 (Surescreen) – achieves point estimates that meet the MHRA criteria. At a threshold of 0.67, we obtain a sensitivity/specificity of (98.4 [CI: 97.1%-99.1%], 98.0 [CI: 96.7%-98.9%]). Interestingly, there is no improvement in predictive accuracy over the two-test combination of tests 6 and 7. In fact, no three-test combination performs better than this two-test combination, in meeting the MHRA’s accuracy criteria.

Adding a third test is of more value in meeting the CDC’s 99.5% specificity criterion. All 14 three-test combinations included in our analysis can achieve an estimated specificity of 99.5%. However, with a three-test combination, the achievable sensitivity is significantly higher. Specifically, combining index tests 2 (Fortress), 6 (Biopanda II), and 8 (Abbott) and applying a threshold of 0.88 (which coincidentally correspond to the MAJORITY heuristic) achieves an estimated sensitivity and specificity of (95.8% [CI: 94.0%-97.1%], 99.6% [CI: 98.7%-99.9%]), with outcome sequences (+, +, +), (+, +, –), (+, –, +), and (–, +, +) classified as positive, and the rest as negative.

### Comparison with benchmark heuristics

While many of the classification thresholds we identify as promising for two-test and three-test combinations correspond to the benchmark heuristics, our results demonstrate the value of exploring other classification schemes. As an example, the ANY, MAJORITY, and ALL heuristics applied to the three-test combination of tests 6 (Biopanda II), 7 (Surescreen), and 8 (Abbott) result in a sensitivity/specificity of (100.0% [CI: 99.7%-100%], 97.4% [CI: 95.9%-98.4%]), (94.8% [CI: 92.8%-96.2%], 99.3% [CI: 98.4%-99.7%]), and (66.5% [CI: 62.8%-70.0%], 99.8% [CI: 99.1%-100%]), respectively. Our threshold approach uncovers 6 other possibilities for classification, including the option (mentioned above) which includes the outcome sequence (-, +, -) in the set of outcome sequences classified as positive. This achieves a sensitivity and specificity of (96.3% [CI: 94.6%-97.5%], 98.9% [CI: 97.8%-99.5%]), which could be optimal for some subpopulations. Note that by applying the thresholds 0.60, 0.94, 0.99 one can obtain sensitivity/specificity values of (96.3% [CI: 94.6%-97.5%], 98.9% [CI: 97.8%-99.5%]), (89.0% [CI: 86.4%-91.2%], 99.6% [CI: 98.7%-99.9%]), and (80.6% [CI: 77.4%-83.5%], 99.8% [CI: 99.0%-100%]), respectively – none of which were identified by the benchmark heuristics.

Our threshold-based classification methodology results in 112 achievable sensitivity/specificity values on the testing possibility frontiers associated with the 14 three-test combinations included in our analysis. While the MAJORITY heuristic represents one point on each frontier, the ANY (ALL) heuristic fails to appear on the frontier for 5 (4) of the 14 combination tests, because is strictly dominated by points on the frontier. Thus, even when the goal is to maximize sensitivity (specificity), for which applying the ANY (ALL) heuristic may seem reasonable, our threshold-based methodology often achieves better results.

## DISCUSSION

We develop and illustrate a methodology for combining diagnostic tests to simultaneously achieve high sensitivity and high specificity, with index tests that are vastly inferior to the best available. The objective of this methodology is not to compare individual index test and their ability to identify various antigen markers, but to combine index tests for better diagnostic accuracy. We use combination positive predictive value (CPPV) estimates to classify each possible outcome sequence of a combination test, accounting for correlated or discordant outcomes.

For any combination test, we are able to evaluate the common ANY, ALL, and MAJORITY heuristics against the ‘testing possibility frontier’ – which represents the trade-off between achievable sensitivity and specificity for that combination. Our approach reveals when benchmark heuristics are inferior. From the ‘envelope’ of frontiers we generate, policy makers can choose points on the envelope – each representing a distinct sensitivity/specificity combination – ideal for particular subpopulations.

Volume discounts can accrue by using the same index tests for different subpopulations, e.g., with a higher sensitivity threshold for older individuals (for whom false negatives can be costly) than for frontline workers (for whom false positives are costly). Strikingly, we observe large gains in accuracy by combining just two or three LFIAs, an approach that is cheaper than a laboratory-based test. The cost of additional tests also needs to be weighed against the cost of a false result - at under &10 per LFIA, offering a three-test kit is likely to be far less costly than a false result.

An LFIA requires little serum, so blood samples can be shared across hospital systems and even countries, to identify the best LFIA combinations. FIND (Foundation for Innovative New Diagnostics, a WHO Collaborating Center) and the CDC are evaluating numerous SARS-CoV-2 immunoassays on standardized samples. Analyzing this data using our methodology would identify the best available combinations of sensitivity and specificity attainable, so that policy-makers can choose optimal combinations for specific populations. This approach would overcome limitations of small sample size and enable more precise parameter estimation. Future research can examine larger samples, and subpopulations with different prevalence rates.

Our methodology also applies when using the same LFIA test multiple times on a sample. Tests with low kappa values are useful candidates for repeat testing as their outcomes have low correlation.

Beyond LFIAs, our threshold-based classification methodology applies to *any* diagnostic tests that can be used in combination, e.g., quick, cheap and inaccurate rapid antigen tests for Covid-19[27], and would enable quick and accurate testing in use cases such as entry to public spaces including airports, nursing homes and hospitals. It can even be used to combine readings from infrared thermometers that rapidly screen for body temperature to permit entrance into buildings.

With our approach, test development is no longer a high stakes winner-take-all game, though clearly continued development of better single tests is desirable. ‘Close enough’ can be good enough – in combination with other tests. Test developers can cooperate and develop complementary tests whose outcomes are less correlated, rather than compete in the same space.

Clinicians often informally combine tests, without considering underlying correlations between test outcomes. Our methodology codifies this practice and provides a pathway that the FDA can use for evaluation and approval for use of combination tests. Home self-testing for SARS-CoV-2 antibodies using LFIAs is feasible.[28] With a pathway in place, combination test kits can be offered for home use, with clear instructions on how to interpret the results, e.g., “read as positive only if both tests give a positive result”. Another option might be to create a single test kit which combines two or three tests “under the hood”, with no extra effort required on the consumer’s part. This would enable the widespread, frequent community testing urgently needed to curb covid-19.

## Data Availability

All data are available upon reasonable request.

## PUBLIC AND PATIENT INVOLVEMENT

It was not appropriate or possible to involve patients or the public in the design, or conduct, or reporting, or dissemination plans of our research.

## ETHICS APPROVAL

The study’s conduct and reporting is fully compliant with the World Medical Association’s Declaration of Helsinki on Ethical Principles for Medical Research Involving Human Subjects. LFIA performance analysis was undertaken as part of the REACT 2 study (refs http://dx.doi.org/10.1136/thoraxjnl-2020-215732 ; https://doi.org/10.1136/bmj.n423), with ethical approval from South Central–Berkshire B Research Ethics Committee (REC ref: 20/SC/0206; IRAS 283805). All participants provided informed consent. Samples for negative controls were taken from the Airwave study approved by North West–Haydock Research Ethics Committee (REC ref: 19/NW/0054).

## ACKNOWLEDGEMENTS

The authors thank Anna Daunt, Anjna Badhan, Jonathan Brown, Rebecca Frise, Ruthiran Kugathasan, and Srishti Katuri for research assistance, and Maitreyee Hazarika and Sadhana Kapur for useful discussions.

## AUTHOR CONTRIBUTIONS STATEMENT

SJ, JJ, JP, and KR conceived, designed and analysed the combination testing methodology we present in this paper. They used data collected in a prior study (Flower, Barnaby, et al. “Clinical and laboratory evaluation of SARS-CoV-2 lateral flow assays for use in a national COVID-19 seroprevalence survey.” Thorax 2020) to illustrate this methodology, and wrote the paper, with interpretation, critical review and feedback from GF, SS, RT, BF, Maya M, Myra M, HA, PE, WB, CA, HW, GC and AD. KR led the research team.

## SUPPLEMENTARY INFORMATION

### SI 1: Index test correlation tables

We examine the correlation of outcomes, true positives, false positives, false negatives, and true negatives between the eight index tests in Tables S1, S2, S3, S4, and S5, respectively

**Table 1:** Index test outcome correlation (p-values in parentheses)

|  | 1 | 2 | 3 | 4 | 5 | 6 | 7 | 8 |
| --- | --- | --- | --- | --- | --- | --- | --- | --- |
|  | Orient | Fortress | Biosure | Biopanda | Wondfo | Biopanda II | Surescreen | Abbott |
| **1** Orient | 1.00 |  |  |  |  |  |  |  |
| **2** Fortress | 0.86<br>(0.00) | 1.00 |  |  |  |  |  |  |
| **3** Biosure | 0.66<br>(0.00) | 0.71<br>(0.00) | 1.00 |  |  |  |  |  |
| **4** Biopanda | 0.69<br>(0.00) | 0.75<br>(0.00) | 0.67<br>(0.00) | 1.00 |  |  |  |  |
| **5** Wondfo | 0.80<br>(0.00) | 0.79<br>(0.00) | 0.63<br>(0.00) | 0.70<br>(0.00) | 1.00 |  |  |  |
| **6** Biopanda II | 0.69<br>(0.00) | 0.76<br>(0.00) | 0.74<br>(0.00) | 0.76<br>(0.00) | 0.57<br>(0.00) | 1.00 |  |  |
| **7** Surescreen | 0.75<br>(0.00) | 0.89<br>(0.00) | 0.72<br>(0.00) | 0.87<br>(0.00) | 0.67<br>(0.00) | 0.77<br>(0.00) | 1.00 |  |
| **8** Abbott | 0.78<br>(0.00) | 0.82<br>(0.00) | 0.73<br>(0.00) | 0.84<br>(0.00) | 0.73<br>(0.00) | 0.79<br>(0.00) | 0.84<br>(0.00) | 1.00 |

**Table 2:** Index test true positive correlation (p-values in parentheses)

|  | 1 | 2 | 3 | 4 | 5 | 6 | 7 | 8 |
| --- | --- | --- | --- | --- | --- | --- | --- | --- |
|  | Orient | Fortress | Biosure | Biopanda | Wondfo | Biopanda II | Surescreen | Abbott |
| 1 Orient | 1.00 |  |  |  |  |  |  |  |
| 2 Fortress | 0.51<br>(0.00) | 1.00 |  |  |  |  |  |  |
| 3 Biosure | 0.54<br>(0.00) | 0.48<br>(0.00) | 1.00 |  |  |  |  |  |
| 4 Biopanda | 0.41<br>(0.00) | 0.50<br>(0.00) | 0.36<br>(0.00) | 1.00 |  |  |  |  |
| 5 Wondfo | 0.75<br>(0.00) | 0.40<br>(0.00) | 0.56<br>(0.00) | 0.49<br>(0.00) | 1.00 |  |  |  |
| 6 Biopanda II | 0.03<br>(0.45) | 0.59<br>(0.00) | 0.19<br>(0.00) | 0.22<br>(0.00) | -0.11<br>(0.00) | 1.00 |  |  |
| 7 Surescreen | 0.03<br>(0.37) | 0.68<br>(0.00) | 0.18<br>(0.00) | 0.23<br>(0.00) | -0.12<br>(0.00) | 0.81<br>(0.00) | 1.00 |  |
| 8 Abbott | 0.03<br>(0.34) | 0.63<br>(0.00) | 0.17<br>(0.00) | 0.23<br>(0.00) | -0.12<br>(0.00) | 0.83<br>(0.00) | 0.88<br>(0.00) | 1.00 |

**Table 3:** Index test false positive correlation (p-values in parentheses)

|  | 1 | 2 | 3 | 4 | 5 | 6 | 7 | 8 |
| --- | --- | --- | --- | --- | --- | --- | --- | --- |
|  | Orient | Fortress | Biosure | Biopanda | Wondfo | Biopanda II | Surescreen | Abbott |
| 1 Orient | 1.00 |  |  |  |  |  |  |  |
| 2 Fortress | 0.27<br>(0.00) | 1.00 |  |  |  |  |  |  |
| 3 Biosure | -0.02<br>(0.62) | 0.16<br>(0.00) | 1.00 |  |  |  |  |  |
| 4 Biopanda | 0.15<br>(0.00) | 0.19<br>(0.00) | 0.14<br>(0.00) | 1.00 |  |  |  |  |
| 5 Wondfo | 0.24<br>(0.00) | -0.01<br>(0.81) | -0.01<br>(0.76) | 0.25<br>(0.00) | 1.00 |  |  |  |
| 6 Biopanda II | 0.09<br>(0.01) | -0.01<br>(0.78) | -0.01<br>(0.72) | -0.01<br>(0.87) | -0.01<br>(0.84) | 1.00 |  |  |
| 7 Surescreen | 0.13<br>(0.00) | 0.16<br>(0.00) | -0.01<br>(0.78) | 0.00<br>(0.90) | -0.01<br>(0.88) | 0.37<br>(0.00) | 1.00 |  |
| 8 Abbott | 0.12<br>(0.00) | -0.01<br>(0.81) | -0.01<br>(0.76) | 0.00<br>(0.89) | -0.01<br>(0.86) | 0.33<br>(0.00) | 0.22<br>(0.00) | 1.00 |

**Table 4:** Index test false negative correlation (p-values in parentheses)

|  | 1 | 2 | 3 | 4 | 5 | 6 | 7 | 8 |
| --- | --- | --- | --- | --- | --- | --- | --- | --- |
|  | Orient | Fortress | Biosure | Biopanda | Wondfo | Biopanda II | Surescreen | Abbott |
| 1 Orient | 1.00 |  |  |  |  |  |  |  |
| 2 Fortress | 0.95<br>(0.00) | 1.00 |  |  |  |  |  |  |
| 3 Biosure | 0.93<br>(0.00) | 0.94<br>(0.00) | 1.00 |  |  |  |  |  |
| 4 Biopanda | 0.97<br>(0.00) | 0.96<br>(0.00) | 0.95<br>(0.00) | 1.00 |  |  |  |  |
| 5 Wondfo | 0.97<br>(0.00) | 0.95<br>(0.00) | 0.94<br>(0.00) | 0.98<br>0.00 | 1.00 |  |  |  |
| 6 Biopanda II | 0.81<br>(0.00) | 0.83<br>(0.00) | 0.82<br>(0.00) | 0.84<br>(0.00) | 0.84<br>(0.00) | 1.00 |  |  |
| 7 Surescreen | 0.94<br>(0.00) | 0.96<br>(0.00) | 0.93<br>(0.00) | 0.97<br>(0.00) | 0.96<br>(0.00) | 0.86<br>(0.00) | 1.00 |  |
| 8 Abbott | 0.82<br>(0.00) | 0.84<br>(0.00) | 0.82<br>(0.00) | 0.85<br>(0.00) | 0.85<br>(0.00) | 0.98<br>(0.00) | 0.86<br>(0.00) | 1.00 |

**Table 5:** Index test true negatives correlation (p-values in parentheses)

|  | 1 | 2 | 3 | 4 | 5 | 6 | 7 | 8 |
| --- | --- | --- | --- | --- | --- | --- | --- | --- |
|  | Orient | Fortress | Biosure | Biopanda | Wondfo | Biopanda II | Surescreen | Abbott |
| 1 Orient | 1.00 |  |  |  |  |  |  |  |
| 2 Fortress | 0.29<br>(0.00) | 1.00 |  |  |  |  |  |  |
| 3 Biosure | 0.27<br>(0.00) | 0.18<br>(0.00) | 1.00 |  |  |  |  |  |
| 4 Biopanda | 0.27<br>(0.00) | 0.29<br>(0.00) | 0.50<br>(0.00) | 1.00 |  |  |  |  |
| 5 Wondfo | 0.23<br>(0.00) | 0.21<br>(0.00) | 0.35<br>(0.00) | 0.42<br>0.00 | 1.00 |  |  |  |
| 6 Biopanda II | 0.04<br>(0.22) | 0.08<br>(0.02) | 0.01<br>(0.72) | -0.03<br>(0.40) | -0.03<br>(0.37) | 1.00 |  |  |
| 7 Surescreen | -0.02<br>(0.63) | 0.41<br>(0.00) | -0.04<br>(0.28) | 0.13<br>(0.00) | -0.03<br>(0.45) | 0.08<br>(0.03) | 1.00 |  |
| 8 Abbott | 0.07<br>(0.05) | 0.09<br>(0.01) | -0.03<br>(0.34) | 0.03<br>(0.44) | -0.02<br>(0.51) | 0.10<br>(0.01) | 0.18<br>(0.00) | 1.00 |

### SI 2: Two test combination performance

Table 6 provides statistics for each two test combination that meets the criteria of both tests having been applied to at least 100 antibody negative and 100 antibody positive samples. The panel on the left includes the CPPV estimates for each outcome sequence (along with the number of observations) and the panel on the right describes the possible range of (Sensitivity, Specificity) and the associated threshold values that produce them. All confidence intervals are calculated using the Wilson method, reflecting the uncertainty in a given ratio as a function of the sample size.

**Table 6:** Statistics for all two test combinations

| All outcome sequences and associated CPPVs |  |  |  | All possible threshold values and associated (Sensitivity, Specificity) |  |  |
| --- | --- | --- | --- | --- | --- | --- |
| Test comb. | Outcome sequence | CPPV | N | Threshold range | Sensitivity (95% CI) | Specificity (95% CI) |
| 1&2 | (+, +) | 97.2% (92.1%, 99.0%) | 107 | $0.50 < t \leq 0.97$ | 85.2% (82.3%, 87.8%) | 99.4% (98.4%, 99.8%) |
| | (+, -) | 50.0% (29.9%, 70.1%) | 20 | $0.43 < t \leq 0.50$ | 93.4% (91.2%, 95.1%) | 97.4% (95.9%, 98.4%) |
| | (-, +) | 42.9% (15.8%, 75.0%) | 7 | $0.01 < t \leq 0.43$ | 95.9% (94.1%, 97.2%) | 96.7% (94.9%, 97.8%) |
| | (-, -) | 1.0% (0.4%, 2.3%) | 496 | $0.00 < t \leq 0.01$ | 100.0% (99.4%, 100.0%) | 0.0% (0.0%, 0.6%) |
| 1&3 | (+, +) | 100.0% (94.3%, 100.0%) | 64 | $0.70 < t \leq 1.00$ | 65.3% (61.4%, 69.0%) | 100.0% (99.4%, 100.0%) |
| | (+, -) | 70.0% (54.6%, 81.9%) | 40 | $0.07 < t \leq 0.70$ | 93.9% (91.7%, 95.5%) | 97.6% (96.1%, 98.6%) |
| | (-, +) | 6.7% (1.2%, 29.8%) | 15 | $0.01 < t \leq 0.07$ | 94.9% (92.8%, 96.4%) | 94.9% (92.8%, 96.4%) |
| | (-, -) | 1.0% (0.4%, 2.4%) | 486 | $0.00 < t \leq 0.01$ | 100.0% (99.4%, 100.0%) | 0.0% (0.0%, 0.6%) |
| 1&4 | (+, +) | 98.1% (90.1%, 99.7%) | 53 | $0.76 < t \leq 0.98$ | 55.9% (51.9%, 59.8%) | 99.8% (99.0%, 100.0%) |
| | (+, -) | 76.1% (62.1%, 86.1%) | 46 | $0.01 < t \leq 0.76$ | 93.5% (91.3%, 95.3%) | 97.6% (96.1%, 98.6%) |
| | (-, -) | 1.2% (0.6%, 2.6%) | 500 | $0.00 < t \leq 0.01$ | 100.0% (99.4%, 100.0%) | 0.2% (0.0%, 1.0%) |
| | (-, +) | 0.0% (0.0%, 79.3%) | 1 | $0.00 < t \leq 0.00$ | 100.0% (99.4%, 100.0%) | 0.0% (0.0%, 0.6%) |
| 1&5 | (+, +) | 97.3% (90.5%, 99.2%) | 73 | $0.62 < t \leq 0.97$ | 76.3% (72.8%, 79.6%) | 99.6% (98.7%, 99.9%) |
| | (+, -) | 61.5% (42.5%, 77.6%) | 26 | $0.50 < t \leq 0.62$ | 93.5% (91.3%, 95.3%) | 97.6% (96.1%, 98.6%) |
| | (-, +) | 50.0% (18.8%, 81.2%) | 6 | $0.01 < t \leq 0.50$ | 96.8% (95.0%, 97.9%) | 97.0% (95.4%, 98.1%) |
| | (-, -) | 0.6% (0.2%, 1.8%) | 495 | $0.00 < t \leq 0.01$ | 100.0% (99.4%, 100.0%) | 0.0% (0.0%, 0.6%) |
| 2&3 | (+, +) | 97.9% (92.6%, 99.4%) | 94 | $0.84 < t \leq 0.98$ | 65.7% (62.0%, 69.3%) | 99.6% (98.8%, 99.9%) |
| | (+, -) | 83.8% (68.9%, 92.3%) | 37 | $0.41 < t \leq 0.84$ | 87.9% (85.1%, 90.1%) | 98.4% (97.2%, 99.1%) |
| | (-, +) | 40.9% (23.3%, 61.3%) | 22 | $0.02 < t \leq 0.41$ | 94.3% (92.2%, 95.8%) | 95.9% (94.1%, 97.2%) |
| | (-, -) | 1.6% (0.8%, 3.1%) | 498 | $0.00 < t \leq 0.02$ | 100.0% (99.4%, 100.0%) | 0.0% (0.0%, 0.6%) |
| 2&4 | (+, +) | 99.0% (94.3%, 99.8%) | 96 | $0.85 < t \leq 0.99$ | 61.7% (57.9%, 65.3%) | 99.8% (99.1%, 100.0%) |
| | (+, -) | 85.1% (72.3%, 92.6%) | 47 | $0.71 < t \leq 0.85$ | 87.7% (84.9%, 90.0%) | 98.4% (97.2%, 99.1%) |
| | (-, +) | 71.4% (35.9%, 91.8%) | 7 | $0.03 < t \leq 0.71$ | 90.9% (88.5%, 92.9%) | 98.0% (96.7%, 98.8%) |
| | (-, -) | 2.7% (1.6%, 4.5%) | 513 | $0.00 < t \leq 0.03$ | 100.0% (99.4%, 100.0%) | 0.0% (0.0%, 0.6%) |
| 2&5 | (+, +) | 100.0% (94.7%, 100.0%) | 68 | $0.65 < t \leq 1.00$ | 73.1% (69.4%, 76.5%) | 100.0% (99.4%, 100.0%) |
| | (+, -) | 65.0% (43.3%, 81.9%) | 20 | $0.55 < t \leq 0.65$ | 87.1% (84.2%, 89.5%) | 98.6% (97.3%, 99.3%) |
| | (-, +) | 54.5% (28.0%, 78.7%) | 11 | $0.01 < t \leq 0.55$ | 93.5% (91.3%, 95.3%) | 97.6% (96.1%, 98.6%) |
| | (-, -) | 1.2% (0.5%, 2.6%) | 501 | $0.00 < t \leq 0.01$ | 100.0% (99.4%, 100.0%) | 0.0% (0.0%, 0.6%) |

Table 6 (continued): Statistics for all two test combinations
|  |  |  |  |  |  |  |
| --- | --- | --- | --- | --- | --- | --- |
| 2&6 | (+, +) | 100.0% (97.3%, 100.0%) | 140 | 0.81 < t <= 1.00 | 73.7% (70.2%, 76.9%) | 100.0% (99.4%, 100.0%) |
|  | (+, -) | 80.6% (65.0%, 90.2%) | 36 | 0.71 < t <= 0.81 | 88.9% (86.3%, 91.1%) | 98.5% (97.2%, 99.2%) |
|  | (-, +) | 70.8% (50.8%, 85.1%) | 24 | 0.01 < t <= 0.71 | 97.9% (96.5%, 98.7%) | 96.9% (95.3%, 98.0%) |
|  | (-, -) | 0.9% (0.3%, 2.3%) | 449 | 0.00 < t <= 0.01 | 100.0% (99.4%, 100.0%) | 0.0% (0.0%, 0.6%) |
| 2&7 | (+, +) | 99.4% (96.5%, 99.9%) | 160 | 0.75 < t <= 0.99 | 83.7% (80.8%, 86.2%) | 99.8% (99.1%, 100.0%) |
|  | (-, +) | 75.0% (46.8%, 91.1%) | 12 | 0.59 < t <= 0.75 | 88.4% (85.8%, 90.6%) | 99.2% (98.2%, 99.7%) |
|  | (+, -) | 58.8% (36.0%, 78.4%) | 17 | 0.02 < t <= 0.59 | 93.7% (91.6%, 95.3%) | 97.8% (96.5%, 98.7%) |
|  | (-, -) | 2.4% (1.4%, 4.1%) | 510 | 0.00 < t <= 0.02 | 100.0% (99.5%, 100.0%) | 0.0% (0.0%, 0.5%) |
| 2&8 | (+, +) | 100.0% (97.6%, 100.0%) | 154 | 0.78 < t <= 1.00 | 81.1% (77.9%, 83.9%) | 100.0% (99.4%, 100.0%) |
|  | (-, +) | 78.3% (58.1%, 90.3%) | 23 | 0.68 < t <= 0.78 | 90.5% (88.0%, 92.5%) | 98.9% (97.8%, 99.5%) |
|  | (+, -) | 68.2% (47.3%, 83.6%) | 22 | 0.01 < t <= 0.68 | 98.4% (97.1%, 99.1%) | 97.4% (95.9%, 98.4%) |
|  | (-, -) | 0.7% (0.2%, 1.9%) | 450 | 0.00 < t <= 0.01 | 100.0% (99.4%, 100.0%) | 0.0% (0.0%, 0.6%) |
| 3&4 | (+, +) | 97.8% (88.4%, 99.6%) | 45 | 0.89 < t <= 0.98 | 47.3% (43.3%, 51.3%) | 99.8% (99.0%, 100.0%) |
|  | (-, +) | 88.9% (56.5%, 98.0%) | 9 | 0.57 < t <= 0.89 | 55.9% (51.9%, 59.8%) | 99.6% (98.7%, 99.9%) |
|  | (+, -) | 56.7% (39.2%, 72.6%) | 30 | 0.05 < t <= 0.57 | 74.2% (70.5%, 77.5%) | 97.0% (95.4%, 98.1%) |
|  | (-, -) | 4.7% (3.1%, 6.8%) | 516 | 0.00 < t <= 0.05 | 100.0% (99.4%, 100.0%) | 0.0% (0.0%, 0.6%) |
| 3&5 | (+, +) | 100.0% (93.1%, 100.0%) | 52 | 0.81 < t <= 1.00 | 55.9% (51.9%, 59.8%) | 100.0% (99.4%, 100.0%) |
|  | (-, +) | 81.5% (63.3%, 91.8%) | 27 | 0.39 < t <= 0.81 | 79.6% (76.2%, 82.6%) | 99.0% (97.9%, 99.5%) |
|  | (+, -) | 39.1% (22.2%, 59.2%) | 23 | 0.02 < t <= 0.39 | 89.2% (86.5%, 91.5%) | 96.3% (94.4%, 97.5%) |
|  | (-, -) | 2.0% (1.1%, 3.7%) | 498 | 0.00 < t <= 0.02 | 100.0% (99.4%, 100.0%) | 0.0% (0.0%, 0.6%) |
| 4&5 | (+, +) | 97.9% (89.1%, 99.6%) | 48 | 0.87 < t <= 0.98 | 50.5% (46.5%, 54.5%) | 99.8% (99.0%, 100.0%) |
|  | (-, +) | 87.1% (71.1%, 94.9%) | 31 | 0.83 < t <= 0.87 | 79.6% (76.2%, 82.6%) | 99.0% (97.9%, 99.5%) |
|  | (+, -) | 83.3% (43.6%, 97.0%) | 6 | 0.03 < t <= 0.83 | 84.9% (81.9%, 87.6%) | 98.8% (97.6%, 99.4%) |
|  | (-, -) | 2.7% (1.6%, 4.5%) | 515 | 0.00 < t <= 0.03 | 100.0% (99.4%, 100.0%) | 0.0% (0.0%, 0.6%) |
| 6&7 | (+, +) | 98.6% (94.9%, 99.6%) | 140 | 0.94 < t <= 0.99 | 72.3% (68.7%, 75.6%) | 99.6% (98.7%, 99.9%) |
|  | (-, +) | 93.8% (79.9%, 98.3%) | 32 | 0.80 < t <= 0.94 | 88.0% (85.2%, 90.2%) | 99.1% (98.1%, 99.6%) |
|  | (+, -) | 80.0% (60.9%, 91.1%) | 25 | 0.01 < t <= 0.80 | 98.4% (97.2%, 99.1%) | 98.0% (96.7%, 98.9%) |
|  | (-, -) | 0.7% (0.2%, 1.9%) | 453 | 0.00 < t <= 0.01 | 100.0% (99.4%, 100.0%) | 0.0% (0.0%, 0.6%) |
| 6&8 | (+, +) | 98.6% (95.1%, 99.6%) | 145 | 0.91 < t <= 0.99 | 74.9% (71.4%, 78.1%) | 99.6% (98.7%, 99.9%) |
|  | (-, +) | 90.9% (76.4%, 96.9%) | 33 | 0.75 < t <= 0.91 | 90.6% (88.1%, 92.6%) | 98.9% (97.8%, 99.5%) |
|  | (+, -) | 75.0% (53.1%, 88.8%) | 20 | 0.01 < t <= 0.75 | 98.4% (97.2%, 99.1%) | 97.8% (96.4%, 98.7%) |
|  | (-, -) | 0.7% (0.2%, 1.9%) | 452 | 0.00 < t <= 0.01 | 100.0% (99.4%, 100.0%) | 0.0% (0.0%, 0.6%) |
| 7&8 | (+, +) | 99.4% (96.4%, 99.9%) | 155 | 0.83 < t <= 0.99 | 80.6% (77.4%, 83.5%) | 99.8% (99.0%, 100.0%) |
|  | (-, +) | 82.6% (62.9%, 93.0%) | 23 | 0.82 < t <= 0.83 | 90.6% (88.1%, 92.6%) | 98.9% (97.8%, 99.5%) |
|  | (+, -) | 82.4% (59.0%, 93.8%) | 17 | 0.01 < t <= 0.82 | 97.9% (96.5%, 98.8%) | 98.3% (96.9%, 99.0%) |
|  | (-, -) | 0.9% (0.3%, 2.2%) | 455 | 0.00 < t <= 0.01 | 100.0% (99.4%, 100.0%) | 0.0% (0.0%, 0.6%) |

### SI 3: Three test combination performance

Table 7 provides statistics for each three test combination that meets the criteria of all tests having been applied to at least 100 antibody negative and 100 antibody positive samples. The panel on the left includes the CPPV estimates for each outcome sequence (along with the number of observations) and the panel on the right describes the possible range of (Sensitivity, Specificity) and the associated threshold values that produce them. All confidence intervals are calculated using the Wilson method, reflecting the uncertainty in a given ratio as a function of the sample size.

**Table 7:**
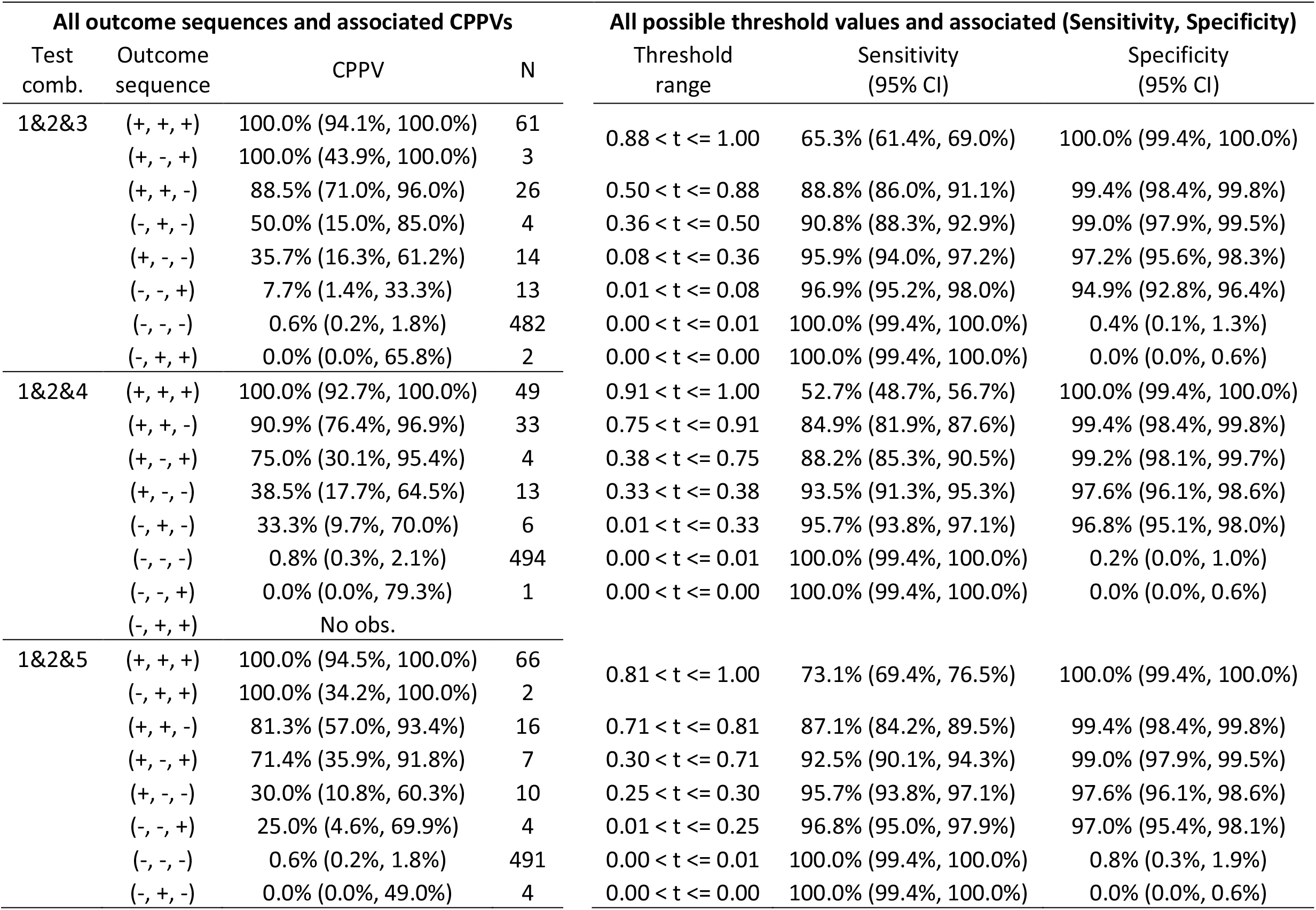
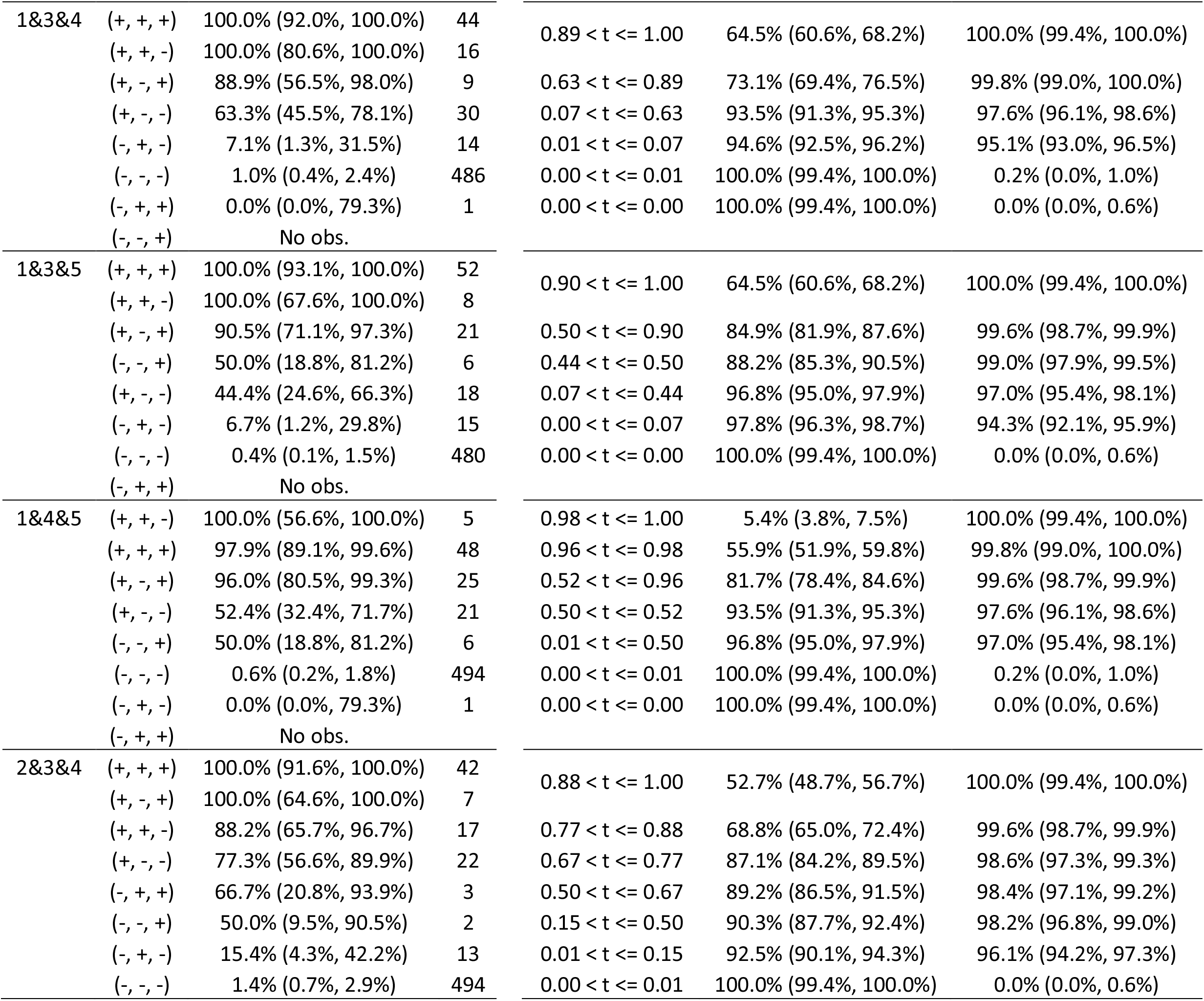
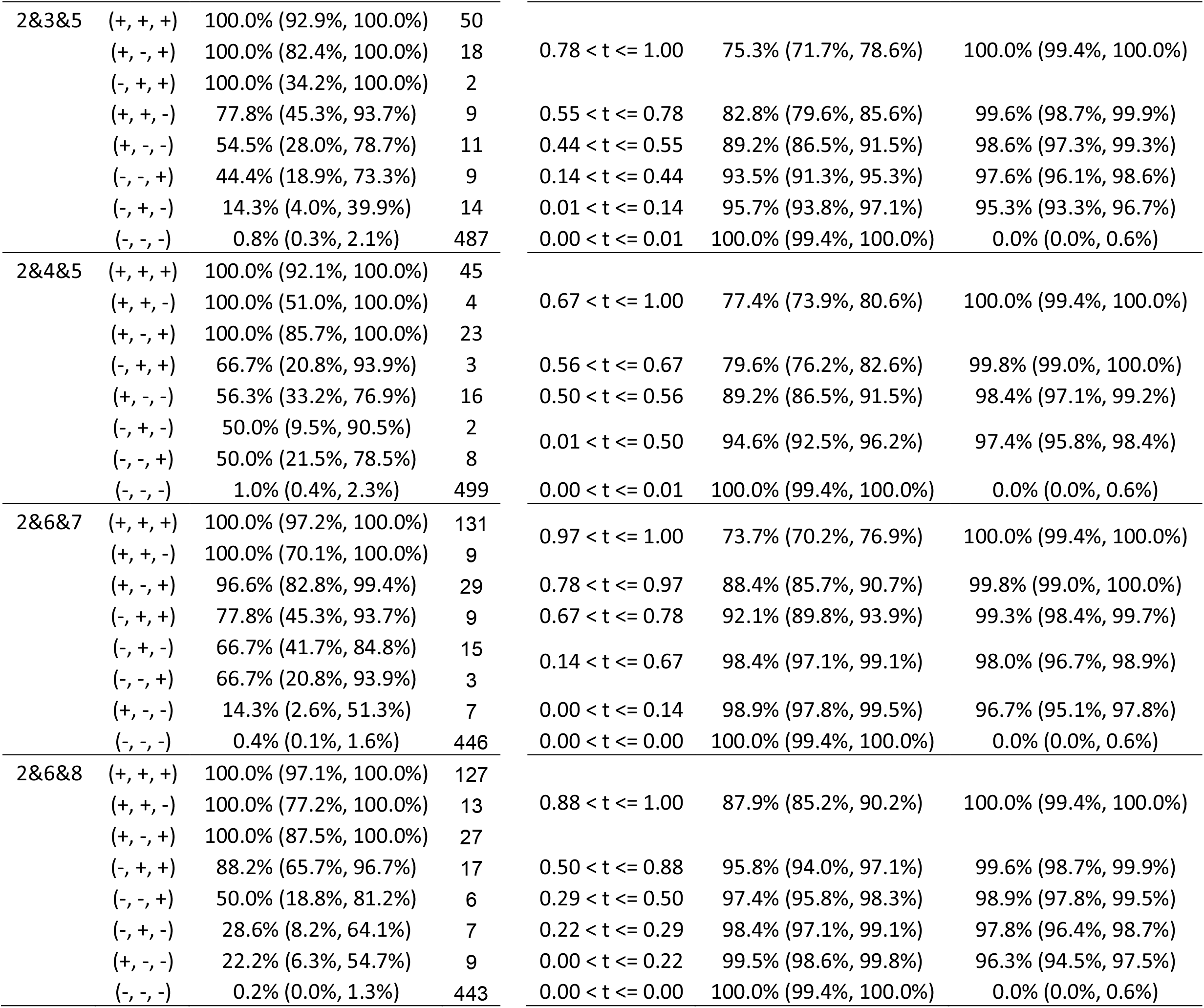
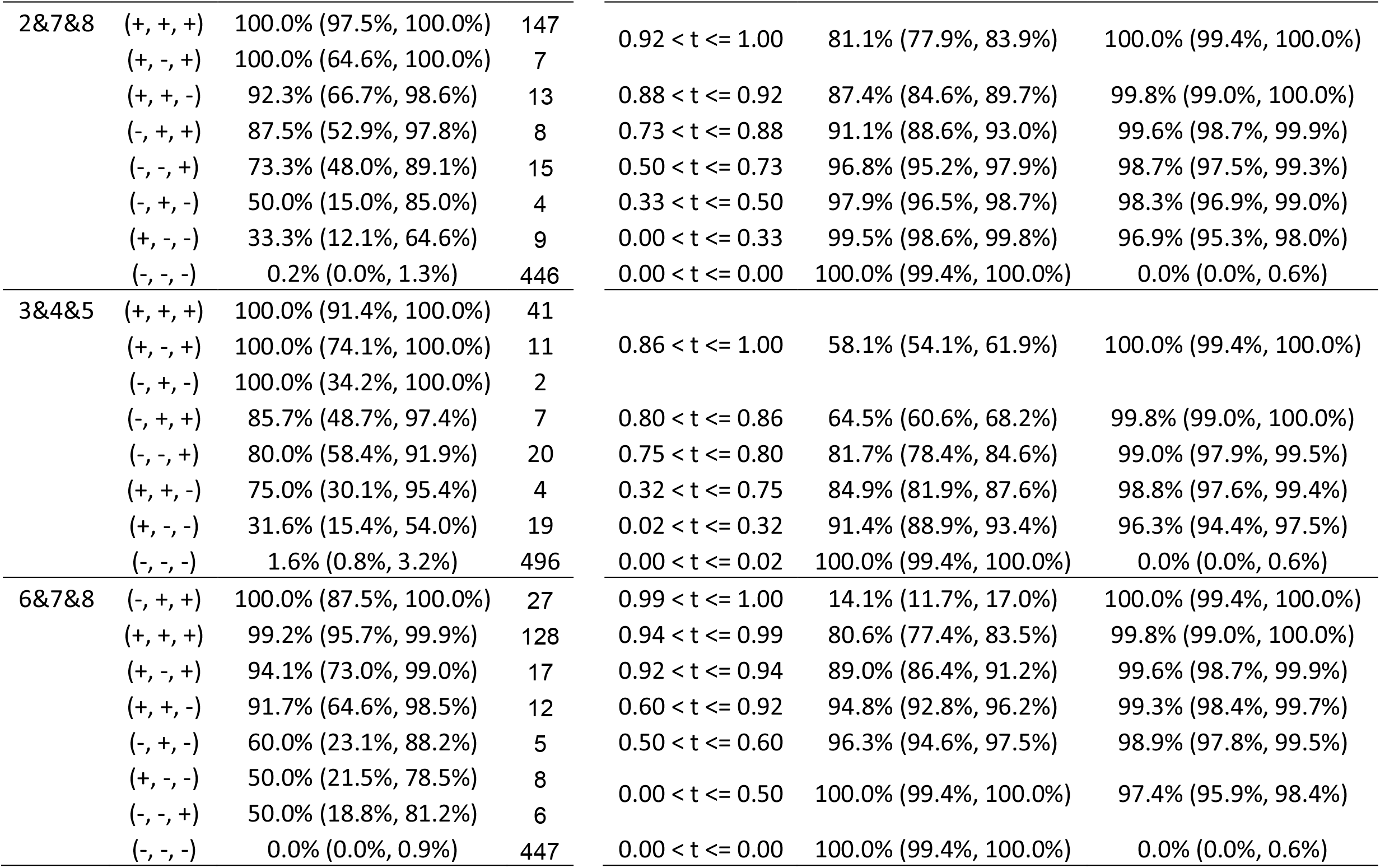
Statistics for all three test combinations

## Footnotes

1* https://www.cdc.gov/coronavirus/2019-ncov/lab/resources/antibody-tests-guidelines.html

2 https://www.cdc.gov/coronavirus/2019-ncov/lab/resources/antibody-tests-guidelines.html

